# Protocol for the development of a tool (INSPECT-IPD) to identify problematic randomised controlled trials when individual participant data are available

**DOI:** 10.64898/2026.02.06.26345217

**Authors:** Calvin Heal, Lisa Bero, George A. Antoniou, Amir Aviram, Nicole Au, Vincenzo Berghella, Esmee M Bordewijk, Paul Bramley, Nicholas J L Brown, Mike Clarke, Lenka Fiala, Steph Grohmann, Lyle C. Gurrin, Jill A Hayden, Kylie E Hunter, Ian Hussey, Brennan C Kahan, Emily Lam, Sarah Lensen, Andreas Lundh, Gideon Meyerowitz-Katz, Ben W Mol, Florian Naudet, Neil E O’Connell, Lisa Parker, Barbara K Redman, Kyle Sheldrick, Emma Sydenham, Madelon van Wely, Rui Wang, Matthias Wjst, Jamie Kirkham, Jack Wilkinson

## Abstract

**Introduction:** Randomised controlled trials (RCTs) investigate the safety and efficacy of interventions. It has become clear however that some RCTs include fabricated data. The INSPECT-SR tool assesses the trustworthiness of RCTs in systematic reviews of healthcare-related interventions. However, where individual participant data (IPD) can be obtained, a more thorough assessment of trustworthiness is possible. Consequently, INSPECT-SR recommends obtaining IPD to resolve uncertainties, though there is no consensus on appropriate methods for forensic analysis of raw data. Our aim is to evaluate IPD checks to establish which are worthwhile, and how they can be implemented in a new tool, INSPECT-IPD (Investigating Problematic Clinical Trials with Individual Participant Data).

**Methods and analysis:** Using international expert consensus and empirical evidence, the INSPECT-IPD tool will be developed in five stages: (1) compiling a list of IPD trustworthiness checks, (2) evaluating the usefulness and ease of interpretation of the checks when applying them to a collection of presumed authentic and fabricated IPD datasets, (3) a Delphi survey to determine which checks are supported by expert consensus, (4) a series of consensus meetings for selection of checks to be included in the draft tool and finally (5) prospective testing of the draft tool in: a) the production of systematic reviews, and b) the journal editorial process for RCT submissions, leading to refinement based on user feedback.

**Ethics and dissemination:** The University of Manchester ethics decision tool determined that ethical approval was not required (18 June 2024). This project includes secondary research and surveys of healthcare researchers on topics relating to their work. All results will be published as preprints and open-access articles, and the final tool will be freely available.

**STRENGTHS AND LIMITATIONS OF THIS STUDY:**

- An international consensus process and empirical evidence will be used to develop the tool.
- The development and dissemination of the tool will involve key stakeholders.
- In the absence of a gold-standard test for problematic data, this tool should not be interpreted as a diagnostic instrument for trustworthiness. Instead, it will assist researchers in assessing the trustworthiness of a study.
- The tool will only be applicable when individual participant data (IPD) can be accessed. Where IPD can be accessed, the ability to assess trustworthiness will be improved.

## Introduction

Randomised controlled trials (RCTs) investigate the safety and efficacy of interventions. Systematic reviews (SR) of health interventions aim to include all RCTs relating to the review question. They combine the results of the RCTs to produce a pooled estimate of the intervention effect and reach an overall conclusion about whether an intervention works and whether it is harmful. The methodological limitations of RCTs are routinely assessed during this process using Risk of Bias (RoB) tools. It has become clear however that some RCTs include fabricated data, and in certain cases, may not have been conducted at all (1). Some false data is a result of research misconduct (including fabricating or falsifying data), while other examples might be due to honest but critical errors. RoB tools are not designed to detect these sorts of problems. We developed the INSPECT-SR (Investigating Problematic Clinical Trials in Systematic Reviews) tool (2), which is designed to assess the trustworthiness of RCTs in systematic reviews of healthcare-related interventions. Through the application of a series of checks the user comes to a judgement on the ‘trustworthiness’ of the study and hence its suitability for inclusion in the SR. Trustworthiness assessment does not depend on the intent of the authors of the studies; its goal is to prevent false data from being included in evidence synthesis. INSPECT-SR guides the user through an assessment based primarily on the information included in study publications and further available documentation, such as the entry in a clinical trial registry. It does not include any forensic examination of the individual participant data (IPD) from the trial. Where IPD can be obtained, a more thorough assessment of trustworthiness is possible.

A recent illustration of the benefits of IPD for the detection of fabricated or otherwise problematic data comes from Carlisle (1). Carlisle assessed 526 RCTs submitted to the journal *Anaesthesia* from February 2017 to March 2020. For trials with no IPD available, he detected false data in 2% and classified 1% as being critically flawed. For trials with IPD available, the detection rate was much higher; he detected false data in 44% of trials and classified 26% as being critically flawed. This strongly suggests that the availability of IPD improves the ability to detect false data, and also suggests that problematic RCTs may be more common than previously thought. Other research has demonstrated the benefits of IPD for detecting integrity problems in a variety of contexts (3). As a result, INSPECT-SR recommends obtaining IPD to resolve uncertainties, mirroring the processes often adopted in research integrity investigations that happen in other contexts (e.g., by journals or publishers). The IPD Integrity tool was developed for detecting integrity issues in randomised trials with IPD available, with items selected for inclusion based on a literature review, consultation with an expert advisory group comprising 13 advisors, piloting and preliminary validation (4,5). Many possible IPD checks were identified during the INSPECT-SR project (6). The current project aims to expand the scope of the work conducted to date by producing empirical evidence for a long list of candidate checks (feasibility, interpretability, performance) and entering them into a large, international consensus process. These various strands of evidence will then be used to select items for inclusion in a new tool acting as an extension to INSPECT-SR when IPD are accessible. The development of this extension will also aim to improve representation of people with expertise and experience in methodology for and conduct of integrity investigations using IPD (for example, undertaking misconduct investigations for journals) compared to research conducted to date. The aim of the current project is therefore to evaluate and select IPD checks for the assessment of trial trustworthiness, and to integrate these into a feasible and useful tool, INSPECT-IPD (Investigating Problematic Clinical Trials with Individual Participant Data).

## Methods and analysis

### Overview

The INSPECT-IPD tool will be developed using five stages (Figure 1), which closely follows the development of INSPECT-SR (2): (1) compiling a list of IPD trustworthiness checks, (2) evaluating the usefulness and ease of interpretation of the checks when applying them to a collection of presumed authentic and fabricated IPD datasets, (3) a Delphi survey to determine which checks are supported by expert consensus, (4) a series of consensus meetings for selection of checks to be included in the draft tool and finally (5) prospective testing of the draft tool in: a) the production of systematic reviews, and b) the journal editorial process for RCT submissions, leading to refinement based on user feedback. A specific subset of systematic reviews, IPD meta-analyses, may be heavily represented in this process, because the researchers conducting them will already have access to, or be in the process of acquiring, the IPD for each included study, bypassing the largest hurdle to implementing the draft INSPECT-IPD tool. It will nonetheless be useful to consider how often reviewers are able to access IPD for the purpose of following up concerns identified using INSPECT-SR.

**Figure 1.**
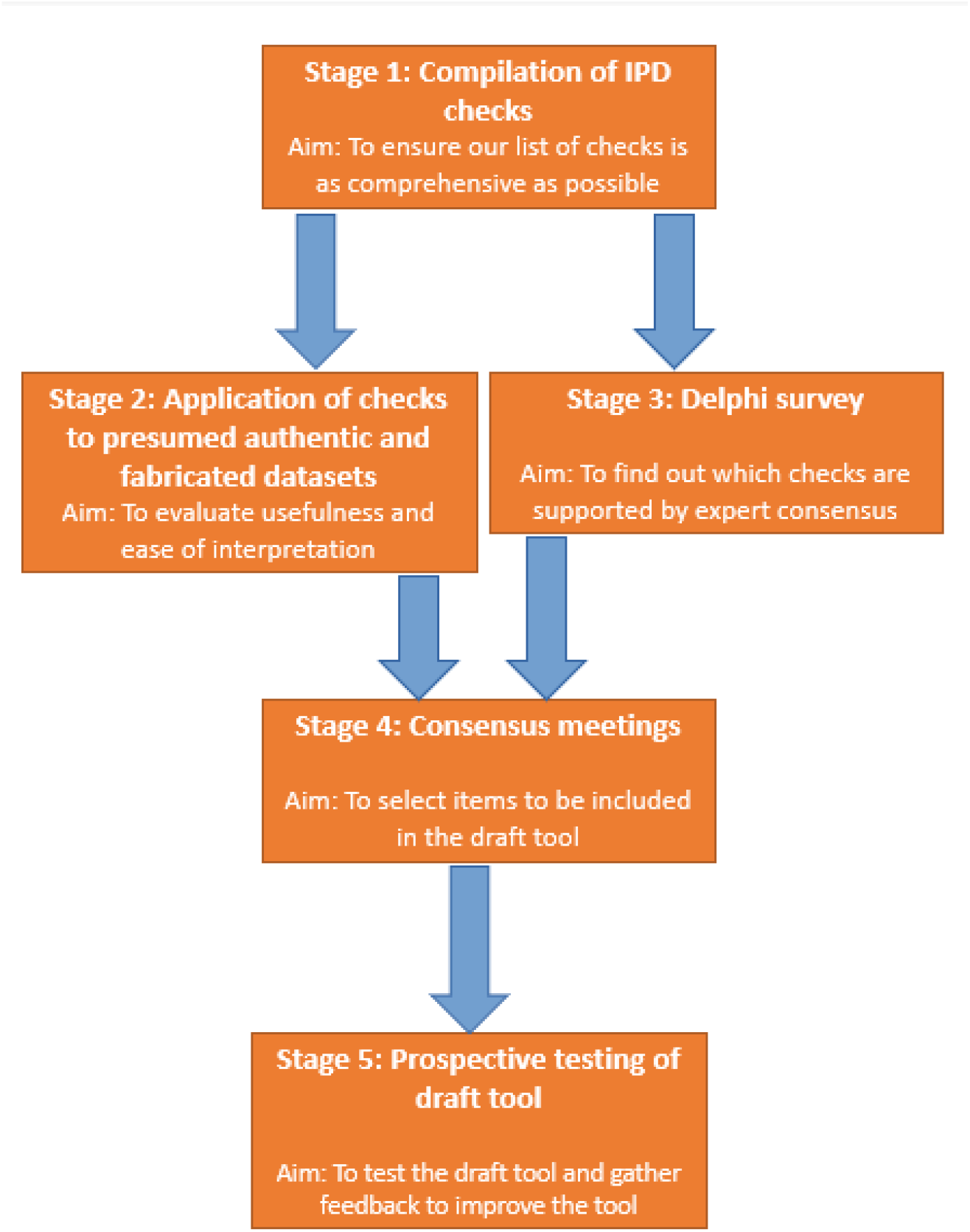
Investigating Problematic Clinical Trials with Individual Participant Data development process.

### INSPECT-IPD working group

The INSPECT-IPD working group comprises a Study Management Group, an IPD Special Interest Group (IPD-SIG), a wider Expert Advisory Panel, a Delphi panel, a lay advisory group and additional collaborators. Individuals can be in more than one of these groups. The IPD-SIG reflects specialist expertise relating to data analysis and forensic analysis of IPD, and will advise on technical aspects of the project. The expert advisory panel has approximately 30 members and was assembled as part of INSPECT-SR but will remain in place throughout INSPECT-IPD, with potential changes to membership according to the availability and identification of people with relevant expertise. This includes research integrity experts, methodologists, researchers with experience in the investigation of potentially problematic trials, systematic review experts, people with clinical trials expertise, journal editors and people representing the interests of patients and lay members

#### Stage 1: Compilation of list of IPD checks

##### Overall design

Stage 1 is complete at the time of submission. An initial list of IPD checks came from Stage 1 of the INSPECT-SR project. These came from a scoping review (7), a qualitative study of experts (8) and additional methods known to the research team. This draft list was presented in a subsequent survey of experts who had the opportunity to comment on these checks and add any not present, forming the initial list used in this study (6). The experts’ feedback on each check was used to improve their wording, as many comments related to their clarity. Identification of further IPD checks was sought via the Expert Panel and IPD-SIG. The expanded list of checks was then classified into domains, with assistance from IPD-SIG and the Expert Panel. The full list of checks and their domains is presented in Supplementary Table 1.

#### Stage 2: Application of the list of checks to presumed authentic and fabricated datasets

##### Overall design

We will apply the full list of checks that we have identified to a sample of presumed authentic datasets and datasets known to be fabricated, in order to establish their usefulness, ease of interpretation and how best to operationalise them. We anticipate that Stage 2 will commence in early 2026 (February-March).

##### Dataset selection

We will collate a sample of 30 datasets, 15 taken from public repositories of trial data and 15 fabricated by the Study Management group using ChatGPT. This method of fabrication has been chosen as large language models are likely to become a common method for fabricating data due to the comparative ease with which they can create simulated data. This sample size has been selected on a pragmatic basis, to allow a sufficient number of applications of the checks to evaluate usefulness and ease of interpretation, while remaining achievable. We will attempt to obtain datasets from trials across a range of health fields.

##### Assessors and recruitment

Stage 2 will be completed as a large collaborative effort, with IPD-SIG members, Expert Advisory Panel members and additional collaborators each applying and/or interpreting all checks in relation to a single IPD dataset. We are aiming for a minimum of 30 assessors to increase diversity in expertise. Each assessor will be assigned an IPD dataset (and its corresponding output) via an allocation list created using random sampling of the datasets without replacement. As we expect to include more than 30 assessors, some IPD datasets may be assessed more than once, and the datasets will be resampled with replacement if this is necessary.

Despite the statistical nature of many of the checks, the intended end-user of INSPECT-IPD will be any researcher completing a systematic review or assessing the trustworthiness of a study. The pool of suitable assessors will therefore not be restricted to individuals with high levels of statistical confidence. The data capture form will record level/area of expertise, allowing us to identify checks that appear to be challenging for people with comparatively less statistical knowledge. If there are checks that are difficult for this audience but which are ultimately determined to be important to include (based on the results of this and the subsequent stages of development), we will attempt to mitigate challenges in interpretation through improved operationalisation and guidance notes in order to include the checks in INSPECT-IPD.

Assessors considered to have made a substantial contribution to data acquisition and critical review of manuscript drafts will be offered co-authorship of the manuscript describing Stage 2.

##### Data capture

Initially the Study Management Team will apply each item from the list of checks from Stage 1 to each of the datasets. The majority of checks will be applied via interaction with an R script designed to produce results relevant to each check. The output of this will be an html file for each dataset which contains the results of each check, including graphs, tables and statistical test outputs, ready for interpretation. The exceptions are those of Domain 1, ‘Inspecting the spreadsheet directly’, which require direct scrutiny of the spreadsheet rather than statistical analysis. Each dataset and accompanying output will be assigned to a minimum of one assessor, though they will remain blinded to the provenance of the datasets.

In the application/interpretation of checks, assessors will be guided by a supplied document briefly explaining the rationale for each check and notes on interpretation. Data collection will be achieved via Excel spreadsheet. For each check, assessors will select a response from ‘not feasible’, ‘not applicable’, ‘no concerns’, ‘some concerns’, ‘serious concerns’ or ‘I don’t understand this check’. This final option allows assessors to indicate that their statistical uncertainty prevents them from answering sensibly. A free-text box will be available for each check so that the assessor may record the reason for their chosen option. Finally, having worked through the list of checks, respondents will record whether they performed any additional checks not included in the list (and if so, what these checks were and what the outcomes were) and give an overall verdict of the trustworthiness of the RCT as a whole, answering the question “Do you have concerns about the trustworthiness of this study?” using one of four response options: ‘no’; ‘some concerns’; ‘serious concerns’; ‘don’t know’. They will also be given the opportunity to make any additional comments and to record how long it took to perform the assessment.

##### Statistical analysis

We will calculate the frequency of each response option for each check (how often each was considered ‘not feasible’ or ‘not applicable’, and how often each one was found to have ‘no’, ‘some concerns’ or ‘serious concerns’). Although we will report the results of each check for each ‘group’ of trials (those presumed authentic and those known to be fabricated), we will not use formal statistical analysis to compare these, as there is no way to truly ascertain the trustworthiness of the trials presumed to be authentic.

#### Stage 3: Delphi survey

##### Overall design

A two-round Delphi survey will be conducted to determine which checks are supported by expert consensus.

##### Participants and recruitment

We will identify Delphi participants through professional networks of the Study Management Team, Expert Advisory Panel, and IPD-SIG. Members of these groups, and members of the INSPECT-SR Delphi Panel, may be eligible to participate. However, the INSPECT-SR project highlighted that many people do not feel comfortable assessing IPD checks, and so it is necessary to supplement the expert population included in the INSPECT-SR Delphi Panel with additional expertise. The INSPECT-SR Delphi Panel included potential users of the tool and those with experience or expertise in assessing potentially problematic studies. We will additionally include individuals with expertise in statistical aspects of RCTs (for example, applied trial statisticians, data managers and clinical epidemiologists), people with experience or expertise in forensic or integrity analysis using IPD, and those who conduct IPD meta-analyses. We will consider 50 expert participants to represent the minimum for a credible Delphi.

##### Selection of items

The list of checks from Stage 1 will form the basis of the list to be assessed in the Delphi survey. The classification and wording of checks may be modified in response to results from Stage 2 or other feedback by the Study Management Group. We will use suitable language to describe the checks, which will be checked for clarity and approved by the special interest group. A written explanation will accompany each check to aid participants in their understanding.

##### Round 1

A personalised email outlining the project will be sent to potential participants. This will include a personalised link to the survey, which will be implemented online using Qualtrics. The survey will include the list of checks. In the first round, respondents will detail their basic demographic and experiential information. Respondents will then score each check from 1 (lowest score) to 9 (highest score) on its usefulness, determined by the perceived potential effectiveness of the check in highlighting untrustworthy data. Participants may also indicate that they feel unable to assess the usefulness of a check, using a ‘Don’t know’ option. Alongside each domain will be a free-text box, allowing participants to leave general comments. We anticipate this may be used by participants to justify their assessment or provide constructive feedback on the wording of the check. First-round participants will also be invited to suggest any additional checks.

##### Round 2

The second round will only include checks for which there was no strong agreement in Round 1; any items that are scored 7 or higher by at least 80% of respondents for usefulness will therefore not be included (but entered automatically into consideration in the consensus meetings). Items that are scored lower than 7 by more than 50% of respondents, however, will not be included in Round 2 nor considered in the consensus meetings. This decision is based on feasibility: with many checks in Round 1, each requiring careful consideration, the burden of survey completion is unavoidably high, so reducing the number of checks in Round 2 is intended to maximise completion rates. Also, in the second round, we will add any additional checks suggested in the first round (subject to review by the IPD-SIG and Expert Advisory Panel). For each item, respondents will be shown both their own scores (1–9) and the distribution of scores from the first round. Potential participants who did not respond to a participation invite to the first round will be invited to the Round 2 survey and will be presented with the distribution of scores from the first round. Participants will be asked to provide their Round 2 score considering this information. The Round 2 survey will include a free-text box for each check so that participants can elaborate on their responses.

##### Analysis

We will summarise the scores from Rounds 1 and 2. Items that are scored 7 or higher by at least 80% of respondents for usefulness, in either round, will be automatically entered into a consensus meeting. Items failing to meet the 80% threshold in Round 2 will be discussed by the Management Group and Expert Advisory Panel and IPD-SIG, and considered for inclusion in the meeting, factoring in the Stage 2 results. We will also summarise the ease of interpretation scores of each check for use in Stage 4.

#### Stage 4: Consensus meetings Overall design

We will hold consensus meetings to finalise the checks to be included in the draft INSPECT-IPD tool. Multiple meetings may be necessary to accommodate international time differences and cover the different domains of the tool. Meetings will be virtual. At the meetings we will present the results of the Stage 2 application exercise and Stage 3 Delphi survey, aiming to finalise the checks to be included in the draft INSPECT-IPD tool. We will consider the ease of interpretation assessments from the Stage 2 application exercise and Stage 3 Delphi as part of this. Some checks may score high for usefulness but low for ease of interpretation; these might be excluded from the main tool but included as an optional or recommended check in the accompanying guidance document. Alternatively, it may be possible to improve ease of interpretation with tailored guidance and formative examples of use. As well as determining the content of the tool, we will determine its form and structure, as well as exactly how its use will interact with that of the INSPECT-SR tool.

#### Stage 5: Prospective testing of draft tool Overall design

In collaboration with systematic reviewers, journal editors, and publishers, we will prospectively evaluate the draft tool by using it in the production of a cohort of systematic reviews and editorial assessments of RCTs submitted to journals.

##### Participants and recruitment

A large network of collaborators representing systematic review groups, journal editors, and publishers has already been assembled for INSPECT-SR, which we will leverage for INSPECT-IPD. Recruited individuals will apply the draft INSPECT-IPD tool while undertaking their systematic review or editorial assessments. Feasibility and usefulness will be assessed by implementing surveys, with separate surveys designed for systematic reviews and editorial assessments. In the systematic review context, the impact on review conclusions will be assessed by comparing results before and after the removal of trials using the tool. Identifying SRs with access to IPD will be a limiting factor. An additional goal is to gain some understanding of the extent of this issue; by the time we come to perform user-testing of INSPECT-IPD, we anticipate that INSPECT-SR will have been in use for around 18 months. This will allow us to identify systematic reviews where there were concerns about the trustworthiness of some of the eligible studies. In these reviews, we will aim to evaluate how often we are able to obtain IPD from the authors of these studies for further investigation, in collaboration with the review authors where possible. A finding that reviewers are frequently unable to obtain IPD would be useful. For testing in the process of undertaking editorial assessments, this may include testing in the conduct of integrity investigations or during the peer review process. Some journals now routinely request IPD for consideration during the editorial assessment process. We believe there will also be value in testing the tool for the purpose of post-publication review of IPD datasets. For example, BMJ now mandates data sharing for published RCTs (9). We will aim to pilot the tool across a variety of potential use-cases and will invite feedback from as many testers as possible within the available timeframe for testing. Stage 5 will culminate in a virtual user workshop. Multiple workshops may be necessary to accommodate international time differences.

##### User workshop

Findings from the surveys will be fed back to participants as part of the user workshop(s). Participants will include systematic reviewers, journal editors, and statistical peer reviewers. Participants will share their experiences of using the tool and make recommendations for refinement. The findings of the testing phase will be used to make final modifications to the tool for usability prior to its completion and publication. We will use the results to produce guidance relating to use of the tool in practice. Alongside Stage 5, as we gather user data, we will produce training materials (workshops and an online training module) to familiarise researchers and editors with the tool.

## Patient and public involvement

Patient partners reviewed and commented on the plans for the project prior to grant submission, and their continued involvement will be sought throughout the study. A PPI panel of eight members has been created for this project. The PPI group will be consulted throughout the project in relation to design of materials, for example, clarity of language in documents, and also in plain language summaries of outputs.

## Ethics and dissemination

The University of Manchester ethics decision tool was used (Supplementary Figure 1). This determined that ethical approval was not required for this project (18 June 2024), as it incorporates only secondary research and surveys of professionals about topics relating to their expertise. Informed consent will be obtained from all survey participants. All results will be published as preprints and open-access articles. The final tool will be freely available.

## Open Science Practices

Each individual stage (from stage 2 onwards) will have their own individual protocols which will be posted on Figshare. Datasets and any analysis code will likewise be stored on Figshare.

## Discussion

Most current attempts at combating untrustworthy RCT data use the summary data reported in the publication, which is frequently all that is available, to judge the trustworthiness of the RCT. However, access to IPD offers an opportunity to more sensitively and completely scrutinise data for trustworthiness. The INSPECT-IPD tool will act as a vital adjunct to the INSPECT-SR tool in the production of systematic reviews, allowing assessors to make judgements about the trustworthiness of RCTs. For journal editors, the tool will present the opportunity to ‘screen’ trials at the pre-publication/reviewing stage and also can be used for post-publication investigations. Optimistically, the availability of tools shown to effectively detect untrustworthy data may make journals more likely to request IPD, and discourage researchers from creating untrustworthy data, leading to the adoption of wider open research practices. Evidence that such a shift has already begun can be found in the data sharing statement by the International Committee of Medical Journal Editors (10), and more recently in policies from both the BMJ (9) and the National Institutes of Health (11), which point towards a near-future of mandatory data-sharing for clinical trials in many journals.

As with INSPECT-SR previously, the development of INSPECT-IPD will benefit from a robust, international consensus process. Key stakeholders including trialists, systematic reviewers, journal editors and patient partners will be involved at all stages. The collaborative nature of the tool’s development will facilitate its effective dissemination, alongside the production of inclusive training materials. In the absence of a gold standard for problematic data, INSPECT-IPD will not act as a diagnostic test for fraud, but rather as a useful tool to determine where studies are sufficiently trustworthy to be included in an evidence synthesis. In applying the checks, those with relevant knowledge of the clinical area and some statistical capability will be helped as they seek a holistic conclusion as to the likely trustworthiness of the data, potentially increasing the trustworthiness of the study conclusion.

A limitation of this study is that despite efforts to incorporate data fabricated by LLM into the tool, the nature of such fraud may evolve in unexpected ways alongside the technology, requiring updates to the tool that reflect this.

## Data Availability

See 'Open Sciences Practices' paragraph

## Declarations

JW declares funding from NIHR for the INSPECT-SR (NIHR203568) and INSPECT-IPD projects (NIHR303741). He also declares statistics or methodological editor roles for BJOG, Cochrane Gynaecology and Fertility, Reproduction and Fertility, Journal of Hypertension. He performs integrity investigations for various journals and publishers. CH declares funding from NIHR for the INSPECT-IPD project (NIHR303741). NOC is a member of the Cochrane Central Editorial Board. Between 2020 and 2023 NOC was Co-ordinating Editor of the Cochrane Pain, Palliative and Supportive Care group, whose activities were funded by an infrastructure grant from the UK National Institute of Health and Care Research (NIHR). He has received funding from the Federal Ministry of Education and Research, Germany under the ERA-NET Neuron Co-Fund Scheme. MC is Co-ordinating Editor for the Journal of Evidence Based Medicine, JLL Library and Cochrane Methodology Review Group. He is one of the founding co-convenors of the Cochrane Individual

Participant Data Meta-analysis (IPD MA) Methods Group and has worked on (and continues to work on) multiple IPD MA and randomised trials. VB is EiC for AJOG MFM, and associate editor for AJOG. LB is Senior Research Integrity Editor, Cochrane, for which she receives remuneration. ES is a Senior Editor of the Cochrane Database of Systematic Reviews. Previously, through March 2023, she was a Co-ordinating Editor of the Cochrane Injuries Group, whose activities were funded by an infrastructure grant from the UK NIHR. She contributed to the Cochrane Editorial Policy on Managing Potentially Problematic Studies. SL is an editor for the Cochrane Gynaecology and Fertility Group, Fertility and Sterility, Human Reproduction Open, and Trials. FN received funding from the French National Research Agency, the French ministry of health and the French ministry of research. He is a work package leader in the OSIRIS project (Open Science to Increase Reproducibility in Science grant agreement No. 101094725). He is also work package leader for the doctoral network MSCA-DN SHARE-CTD (HORIZON-MSCA-2022-DN-01 101120360), funded by the EU. He serves as an academic editor for PLOS One and as an associate editor for Fundamental and Clinical Pharmacology. AA is the trustworthiness editor for AJOG MFM. LP is on the Editorial Board for Journal of Clinical Epidemiology. Paul Bramley is an editor for the journal ‘Anaesthesia’. AL is on the editorial board of BMC Medical Ethics. Kylie E Hunter led development of the Individual Participant Data (IPD) Integrity Tool (12). Rui Wang is a Deputy Editor of Human Reproduction Update and Editorial Board Member of BJOG and Cochrane Gynaecology and Fertility Group. He is a co-author of the Individual Participant Data (IPD) Integrity Tool. He is supported by an NHMRC Emerging Leadership Investigator grant (2009767). MvW is Editor-in-Chief of Human Reproduction Update and senior Editor of the Cochrane Database of Systematic Reviews and Co-ordinating Editor of the Cochrane Gynaecology and Fertility Group and Sexually Transmitted Infections Group. BWM is an Editor of Fertility Sterility and Editorial Board Member of the Cochrane Gynaecology and Fertility Group. He is a co-author of the Individual Participant Data (IPD) Integrity Tool. He is supported by an NHMRC Investigator grant. He reports consultancy for Ferring, Organon, Repronovo, UNILAB and Norgine. BKR, MW, NJLB, EL, IH, EMB, SG, LG, GMK, BK, JAH declare no conflict of interest.

## References

1. Carlisle JB. False individual patient data and zombie randomised controlled trials submitted to Anaesthesia. Anaesthesia. 2021 Apr;76(4):472–9.

2. Wilkinson J, Heal C, Antoniou GA, Flemyng E, Alfirevic Z, Avenell A, et al. Protocol for the development of a tool (INSPECT-SR) to identify problematic randomised controlled trials in systematic reviews of health interventions. BMJ Open. 2024 Mar;14(3):e084164.

3. Lawrence JM, Meyerowitz-Katz G, Heathers JAJ, Brown NJL, Sheldrick KA. The lesson of ivermectin: meta-analyses based on summary data alone are inherently unreliable. Nat Med. 2021 Nov;27(11):1853–4.

4. Hunter KE, Aberoumand M, Libesman S, Sotiropoulos JX, Williams JG, Aagerup J, et al. The Individual Participant Data Integrity Tool for assessing the integrity of randomised trials. Research Synthesis Methods. 2024 Nov;15(6):917–39.

5. Hunter K, Aberoumand M, Libesman S, Sotiropoulos J, Williams J, Li W, et al. Development of the Individual Participant Data (IPD) Integrity Tool for assessing the integrity of randomised trials using individual participant data [Internet]. 2023 [cited 2024 Jun 13]. Available from: http://medrxiv.org/lookup/doi/10.1101/2023.12.11.23299797

6. Wilkinson J, Heal C, Antoniou GA, Flemyng E, Avenell A, Barbour V, et al. A survey of experts to identify methods to detect problematic studies: stage 1 of the INveStigating ProblEmatic Clinical Trials in Systematic Reviews project. Journal of Clinical Epidemiology. 2024 Nov;175:111512.

7. Bordewijk EM, Li W, Van Eekelen R, Wang R, Showell M, Mol BW, et al. Methods to assess research misconduct in health-related research: A scoping review. Journal of Clinical Epidemiology. 2021 Aug;136:189–202.

8. Parker L, Boughton S, Lawrence R, Bero L. Experts identified warning signs of fraudulent research: a qualitative study to inform a screening tool. Journal of Clinical Epidemiology. 2022 Nov;151:1–17.

9. Loder E, Macdonald H, Bloom T, Abbasi K. Mandatory data and code sharing for research published by The BMJ. BMJ. 2024 Mar 5;q324.

10. Taichman DB, Sahni P, Pinborg A, Peiperl L, Laine C, James A, et al. Data Sharing Statements for Clinical Trials: A Requirement of the International Committee of Medical Journal Editors. Ann Intern Med. 2017 Jul 4;167(1):63–5.

11. Ross JS, Waldstreicher J, Krumholz HM. Data Sharing — A New Era for Research Funded by the U.S. Government. N Engl J Med. 2023 Dec 28;389(26):2408–10.

12. Hunter KE, Aberoumand M, Libesman S, Sotiropoulos JX, Williams JG, Aagerup J, et al. The Individual Participant Data Integrity Tool for assessing the integrity of randomised trials. Research Synthesis Methods. 2024 Nov;15(6):917–39.

