## Supplementary material for "Protocol for the development of a tool (INSPECT-IPD) to identify problematic randomised controlled trials when individual participant data are available"

Supplementary Table 1. Initial list of trustworthiness checks, arranged in preliminary domains

**Domain 1 – Inspecting the spreadsheet directly (11 checks)**

| **#** | **Check** | **Description** |
| --- | --- | --- |
| 1 | Does comparing multiple versions of a spreadsheet containing study data reveal issues with consistency? | Discrepancies between spreadsheets which should be consistent may signal issues with authenticity (1). |
| 2 | Does examining the spreadsheet reveal any formulae used to fabricate data? | Examine the spreadsheet for formulae used to generate false data. For example, a formula in the column for patient weight, such as (35kg + (height in metres times a random number between 10 and 30), would be a clear indicator of fabrication. On the other hand, a column representing BMI would be expected to contain the formula to calculate this from height and weight. Indeed, the absence of such a formula could be considered questionable as it would imply, at a minimum, that BMI had been calculated elsewhere and copy/pasted, with a risk of loss of precision. Excel has an option to allow all formulae to be displayed instead of the value that each formula produces in any given cell. |
| 3 | Does changing the global number format in Excel to ‘general’ reveal anything unusual in relation to the number of decimal places recorded? | Values that have been calculated using one or more other variables may display a large number of digits to the right of the decimal place, while those that have been inputted manually are likely to display fewer digits. Inspecting the numbers of digits can therefore reveal problems, for example, if there is heterogeneity in the number of decimal places recorded across observations within a variable (1,2). |
| 4 | Does examining the ‘calcChain’ of the Excel file reveal reordering of rows after creation or other unexpected manipulation? | Windows 11 instructions   1) Navigate to excel file in file explorer 2) Click ‘View’ in file explorer (not the file), click ‘Show’ and ensure ‘File name extensions’ is ticked 3) Copy and paste the excel file (to create backup) 4) Rename the excel file to end in ‘zip’, so it should now look like “filename.xlsx.zip” 5) Right click, ‘Extract all’, ‘Extract’ 6) In ‘xl’ folder you’ll find calcChain.xml 7) Copy and paste the text within into [Free Online XML Formatter - FreeFormatter.com](https://www.freeformatter.com/xml-formatter.html) and click ‘Format XML to new window’ 8) Examine the code for unnatural ordering or broken sequences, see <https://datacolada.org/109> on how to do this. (3) |
| 5 | Does colour coding values in the spreadsheet highlight outlying values, patterns or repetition? | Colour coding the values in a spreadsheet (for example, using conditional formatting) may make it easier to spot repetition or other signs of fabrication (4). |
| 6 | Does checking the meta-data in the sheet (when it was created, by whom, number of hours it’s been opened) reveal anything contradictory to the publication? | The proposal is to view the meta-data from within the excel file by using ‘File’ → ‘Info’. This may reveal information incompatible with what has been reported, such as dates that don’t align. Many caveats apply here; caution is required. For example: copying/moving files can reset "Created" date to copy date. |
| 7 | Does reordering rows by different variables reveal previously unnoticeable patterns? | Sorting by variables within excel, perhaps in conjunction with the colour coding method above, may reveal patterns unexpected of genuine data. |
| 8 | Are there any data fields missing from the IPD? | Check that the variables included in the manuscript are included in the dataset. A caveat though: it may be standard protocol in some contexts (i.e. with CTU involvement) for only requested variables to be included. |
| 9 | Are there formatting artefacts? | Authentic data may be more likely to contain consistent and logical formatting (2). The proposal is to check for odd patterns of formatting. An example may be two different types of text font used, which may signal that some data has come from a separate document. The suspiciousness of any such artefacts may depend on the details; for example, a whole column being formatted differently is unlikely to be suspicious, but individual cells within a column being formatted differently may be considered suspicious (2). |
| 10 | Are there indications content has been imported from statistical software? | Statistical software will often truncate the content of string variables if they exceed a certain number of characters. This may be indicated if multiple values of a string variable are an arbitrary value. For example, if 80 characters appears to be the limit the data may have been imported from Stata (where 80 characters is the string limit). |
| 11 | Does the file format used to distribute the data make sense? | Data are typically analysed with software. While most proprietary packages can read .CSV or .XLS[X] files, it would be more common for a dataset from SPSS to be in .SAV format, from Stata in .DTA format, etc. Sometimes authors might export data to a more open format to reduce the dependence of subsequent analysts on their software, but in other cases the fact that the data are only available as .CSV or (especially) .XLS[X] format might indicate that this is not the file from which the analyses were run. This is especially the case if the file contains any evidence of having been entered manually (see previous checks). |

**Domain 2 – Inspecting sequences (8 checks)**

| **#** | **Check** | **Explanation** |
| --- | --- | --- |
| 12 | Does plotting variable values in the order provided by the author, and then by study group, reveal any repetition, patterns or differences in patterns between groups? | **On application to outcome data:** Systematic differences between study groups at outcome may be expected, but repetitions or patterns, especially across several variables, may signal untrustworthy data. Or rather, the proposal is to check this graphically.  **On application to baseline data:** Under genuine randomisation we would expect independence of observations, this would lead to randomness in sequential values where few repetitions or patterns would arise. The proposal is to check this graphically. Some caution is required; there may be genuine reasons for patterns (i.e. seasonal changes for data collected over a long time period). |
| 13 | Does plotting differences between consecutive values within a variable in the order provided by the author and then by study group reveal any repetition, patterns or differences in patterns between groups? | Similar to the above check but instead of plotting raw values, plotting instead the difference between consecutive values, which may identify cases where a fabricator has copied and edited the copied values. |
| 14 | Does estimating the probability of column sequences via simulation and resampling reveal irregularities? | One method of fabrication may be copying columns of data from one group and pasting them to another, making small changes to the pasted values. If this has occurred, we will end up with small differences between groups when comparing their sequential rows (i.e. the first patient in group 1 will have similar values to the first patient in group 2, and so on for the subsequent patients). Graphical assessment illustrates this, but simulation/resampling allows us to assign a probability of such similarities occurring by chance. |
| 15 | Are tests of runs within a variable suggestive of fabrication? | The randomness of runs in a variable can be tested. Fabricators may be poor random number generators and may change values too frequently or too infrequently compared to what would be expected in authentic data. This can be applied to continuous variables by testing runs of values occurring above/ below some threshold value. |
| 16 | Are there implausible sequences in decimal places (after deleting integer)? | Some fabricators may copy and paste values and change the integer portion to disguise this. Plotting the data without the integer portion may reveal patterns in such a case (4). |
| 17 | Are there implausible sequences in integers (after deleting decimals)? | Some fabricators may copy and paste values and change the decimal portion to disguise this. Plotting such values without the decimal portion may reveal patterns in such a case (4). |
| 18 | Is there evidence of autocorrelation between successive observations in the dataset? | In authentic data, we may not expect serial correlation between measurements taken on distinct study participants. By contrast, if a fabricator has entered values into a column manually, there may be signs of serial correlation across rows due to the fact that human beings are typically poor random number generators. Autocorrelation should be examined (for example, graphically) in dataset order and separately for each study group. For baseline variables, autocorrelation should not differ between study groups. Methods exist for testing autocorrelation, e.g. Durbin-Watson method. |
| 19 | When the dataset is ordered by participant ID or randomisation timestamp, does the N+1th participant have the same condition as the Nth 1/k of the time (where there are k conditions)? | The proposal is to check for repeated values of categorical variables occurring every Nth row. An example, with two categories of gender, after every male patient we have a 50% chance of the next patient also being male. If the condition assignment has been fabricated "by hand", the condition will often change too frequently as the faker tries to avoid "excessively long identical sequences”. |

**Domain 3 – Inspecting relationships between variables (11 checks)**

| **#** | **Check** | **Explanation** |
| --- | --- | --- |
| 20 | Can inliers be identified using singular value decomposition? | Inliers are values that skew unnaturally close to the average. An example could be someone with very high BMI who otherwise has average clinical measurements. To investigate this each data element is imputed (as if missing) based on the other data elements in the matrix and then the imputed value is compared to the observed value. If the observed value is much lower than the imputed value, this suggests an inlier (5). |
| 21 | Can inliers be identified using Mahalanobis distance? | Similar to above, inliers can be determined by the presence of lower than expected Mahalanobis distances. |
| 22 | Does examining relationships between variables suggest biological/contextual implausibility? | The proposal is to check if variables that are known to be biologically associated display the expected relationships (e.g. by using a scatter plot). An example would be checking that height and weight are positively correlated. Perfectly correlated variables may also be revealed by this method. Another proposal is to use heatmaps to examine correlations between variables. |
| 23 | Do statistical tests that compare multivariate correlations between variables show differences between study groups? | Multivariate correlation between variables at baseline should be unrelated to randomisation, therefore the proposal is to check that these do not meaningfully differ between the study groups. Tests for comparing covariance matrices may be employed. |
| 24 | Does plotting Chernoff faces for each group reveal any unusual patterns or differences? | Plotting Chernoff’s faces may reveal outliers or unexpected homo/heterogeneity within/between groups. Ordering by different variables might aid the assessment. |
| 25 | Does plotting Star Plots for each group reveal any unusual patterns or differences? | Plotting star plots may reveal outliers or unexpected homo/heterogeneity within/between groups. Ordering by different variables might aid the assessment. |
| 26 | Does applying a neighbourhood clustering method reveal unusual/unexpected clustering? | One way to fabricate data is make observations similar to that of genuinely recorded data. If this has occurred there may be more clustering of data than would naturally arise. Several statistical models serve to highlight such clustering, with the aid of graphical outputs (6). |
| 27 | Are any variables a subset of a second variable within a data set? | Does it appear as though the values of variables may have been copied and pasted from other variables (with some tweaks)? This is similar to the check: “Does estimating the probability of column sequences via simulation and resampling reveal irregularities?”, however where that check was comparing values between groups (i.e. is age in group 1 too similar to age in group 2), this check is comparing variables across the whole dataset (i.e. is outcome 1 too similar to outcome 2). This check may flag variables that are highly correlated or structurally dependent (e.g. live births are a subset of pregnancies) and so a positive result is only suspicious if you suspect the highlighted variables should not naturally be correlated. |
| 28 | Is there evidence of abnormally low within person variability? | It is biologically/clinically expected that certain measures should change over time. The proposal is to consider the recorded stability of such measures. |
| 29 | Is there evidence within repeated measures of interpolation and duplication? | Repeated measures data may be fabricated from recorded, genuine measurements (e.g., copying a patient’s time 1 value to time 2, or adding or multiplying by a constant). The proposal is to check for evidence of this, for example by plotting repeated measurements against each other, or using profile plots of longitudinal data (7). |
| 30 | Does applying principal component analysis reveal abnormal loading on the first component? | If several variables are linear transformations of an original variable, they’ll all be measuring the same underlying construct. If PCA reveals abnormally large loading on the first component this may be evidence of this. In genuine trial data, many different constructs would be measured, and none would be suspiciously dominant. |

**Domain 4 – Inspecting digit/variable distribution (17 checks)**

| **#** | **Check** | **Explanation** |
| --- | --- | --- |
| 31 | Does examination of leading digits suggest violation of Benford's Law? | If the magnitude of a variable allows, we might expect the leading and subsequent digits of a continuous variable to follow a pattern described by Benford’s Law, specifically that leading digits will more frequently be smaller (1 or 2 for example) (8). However, this law will not often apply to clinical trial data, which is typically bounded, so caution is merited (9). |
| 32 | Does the distribution of non-first digits deviate from the approximately uniform distribution that is typically expected under Benford's Law? | Under Benford’s Law, the final digit will be expected to be roughly uniformly distributed (8,10) |
| 33 | Do distributions of leading digits in baseline variables differ between randomised arms? | Regardless of the nature of the distribution (i.e., whether it should follow Benford’s Law), under genuine randomisation we would expect the distributions of leading digits in baseline variables to be similar between groups. This can be tested using a chi-squared test (or some variation thereof) (11). |
| 34 | Do distributions of non-leading digits differ between randomised arms? | The proposal is to check the similarity of distributions of non-leading digits between groups. This can be tested using a chi-squared test (or some variation thereof). |
| 35 | Do the distributions of single variables follow simple but implausible models? | The proposal is to check if variables follow common probability distributions (such as Normal) more closely than would be expected in genuine data. A lack of noise may suggest that the values were simulated using statistical software. |
| 36 | Is there a detectable round number preference/excess of round numbers? | An excess of ‘5’ or ‘0’ in the final digits of variables with no contextual justification may be evidence of digit preference, or a ‘rounding up/down’ of values when recording, a potential sign of data fabrication. (12) |
| 37 | Do the frequency of digits suggest violation of Zipf’s law? | Zipf’s law describes how, when a variable is ranked by how frequently each value occurs, each frequency is closely related to its rank. The original usage suggests that “the successive relative populations of a series of quantities are roughly proportional to 1, 1/2, 1/3, 1/4”. Applying this to categorical variables, the proposal is to consider whether this law appears violated (for example, if the second most frequent value of a variable was only marginally less frequent than the most frequent value) (13). |
| 38 | Are there an implausible number of outliers detectable? (using graphical or other methods) | A surplus of outliers may signal data that has been fabricated to achieve a desired result (effect), the proposal is to check for these. |
| 39 | Are unexpected peaks in variable distributions evident? | Many variables follow known distributions to varying degrees. Unexpected peaks may signal duplicated values or digit preference. For baseline values, peaks in one arm but not the other may suggest lack of proper randomization (12). |
| 40 | Are variables skewed beyond expectation? | Taking the expected distribution and sample size into account, the proposal is to consider whether there is excessive skew in the observed distribution of any variables. |
| 41 | Is there too little or too much variance? | The proposal is to check that the amount of variance falls within reasonably expected limits. Fabricators may “underappreciate the higher level sampling fluctuations, resulting in generating too little randomness (i.e., error) in the standard deviations across groups” This can be tested using IPD by comparing the observed vs expected distribution of variances (10). |
| 42 | Is there an unusually low proportion of missing data? | The suggestion here is to consider whether the amount of missing data in the study is plausible given the clinical context. Missingness is normal in authentic trials and its absence may be suspicious (14). |
| 43 | Are there missing participant IDs (if sequential)? | Missing IDs may signal the numbers were fabricated rather than collected, or poor record-keeping. |
| 44 | Do missing data appear too symmetrical across groups or implausibly few? | Frequency of missingness may be naturally expected to vary by study groups, especially at the endpoint. |
| 45 | Are the coefficients of variation implausibly similar between variables and between study groups? | The proposal is to compare the coefficients of variation between variables, firstly within each group and then between groups. Fabricated values may yield coefficients of variation that are more similar between unrelated variables than would normally arise (14). |
| 46 | Is there unexpected variation in the number of decimal places reported within a variable? | The proposal is to examine the frequency of meaningful decimal places in each numeric variable. For example, if a variable is generally recorded to two decimal places, most observations will have two meaningful decimal places (e.g., 3.15), some will have one decimal place (e.g., 4.2), and very few will be integers (e.g., 7). |
| 47 | Is there evidence of ‘number-bunching’ when analysing the frequency with which values get repeated within a dataset. | For variables with multiple digits (e.g. 1.23) the proposal is to assess the 'average frequency' of values by splitting the first from later digits (e.g. 1 then 0.23), and resampling to get a sense of how unusual the provided distribution is. (15) |

**Domain 5 – Inspecting internal/external consistency (12 checks)**

| **#** | **Check** | **Explanation** |
| --- | --- | --- |
| 48 | Is it impossible to reproduce the results in the paper from the underlying dataset? | The proposal is to run statistical tests on the IPD to determine if the results reproduce those in the paper. |
| 49 | Does participant ID number fail to correspond to randomisation order? | If randomisation date or order is recorded, does it correspond to participant ID? I.e., Does participant ID 1 have the earliest recorded randomisation date? |
| 50 | Are there unexplained inconsistencies in overall participant numbers and participant numbers per group? | The proposal is to check that the number of participants in each group match that of the related published documents (e.g. CONSORT diagram, registration record, publication text/tables). |
| 51 | Are data internally inconsistent? | Some results are mutually exclusive. For example having a weight of 50kg and a BMI of 45, or being male and having Turner syndrome. |
| 52 | Does an interaction test to assess the subgroup homogeneity suggest implausible consistency? | The Tarone test determines whether two (or more) odds ratios are homogenous. In some RCTs we might expect subgroups to have different odds ratios (e.g., the association in question might be expected, a priori, to be stronger in men compared to women). A significant test signals evidence against the null of homogeneity. |
| 53 | Are there concerns relating to the frequency or nature of data errors? | The proposal is to check for an excess of errors (for example in the reported units of variables, or typographical errors) that may suggest non-expertise. |
| 54 | Are there discrepancies between IPD (or reported data) and eligibility criteria specified in publication(s) and/or registration record(s)? | The proposal is to check if participant data contradict the reported eligibility criteria, i.e. by comparing the minimum/maximum values of variables and any inclusion/exclusion criteria that refer to these (16). |
| 55 | Are there any other external inconsistencies between publication, registration, IPD? | Generally, are the IPD consistent with the publication/other documents (with regards to, for example: sample size, location, results, study methods)? Previous checks have covered elements of this, so this is somewhat of a catch-all for anything not previously specified, but may include something like ‘do the number of patients at each site match?’ |
| 56 | Is the randomisation sequence inconsistent with the description in the paper? | The possibility of the number of sequential patients allocated to a trial arm can be considered given the method of allocation in the study. For example, with blocked randomisation with block sizes of 4, it would be impossible for 5 patients to sequentially be allocated to the same arm, and we know that 4 patients being allocated to the same arm sequentially should only happen infrequently. |
| 57 | Is there evidence that participants in the dataset may have been duplicated? | The proposal is to check whether rows of data may have been duplicated. This may be full duplication or, more commonly, partial duplication where some values may have been edited to conceal the duplication. One way to achieve this is scanning the dataset for repeated value combinations, starting with the maximum number of variables and gradually decreasing. For example, if a dataset has 'BMI, Age, Ethnicity, Sex', the method will count the frequency of participants who match on all four of these, then look at how many match on three of these, and so on. If certain combinations occur unusually often, this may be an indication of data fabrication, and it might be worth extracting those rows for further inspection (4). |
| 58 | Are there repeating data patterns across rare variables? | Sort any rare variables (i.e., categorical variables with rarely occurring values) by value, looking for patterns. Participants who experience a rare value of a variable and have the same characteristics over other variables may signal duplication. For example, one might find that all patients with a rare blood type have the same date of birth and are the same sex. |
| 59 | Does removal of outliers meaningfully alter the effects of interest? | Fabricators may add a few extreme data points to get the result they want. The proposal is to compare the results when running the analyses with and without the outliers. |

**Domain 6 – Inspecting timings in the dataset (5 checks)**

| **#** | **Check** | **Explanation** |
| --- | --- | --- |
| 60 | Do visit dates align with contextual expectations? Are visit dates uniformly distributed across possible dates across groups? | The proposal is to check whether visit dates fall on unlikely days (e.g., weekends for data collected at out-patient clinics). Caution is required here: international/cultural differences may explain seemingly unlikely recruitment patterns. Also, among plausible visit dates, are numbers of visits reasonably uniform? (17) |
| 61 | Does inspecting recruitment over calendar time between groups highlight unlikely differences? | The proposal is to check for large discrepancies in recruitment time between groups; knowledge of the reported randomization procedure may aid in this. A cumulative incidence frequency plot may be used. |
| 62 | Does inspecting time between participant visits reveal irregular or unexplained intervals? | Given knowledge of expected regularity of visit dates from the publication, the proposal is to determine whether the observed time between visits is plausible. |
| 63 | Are differences between actual visit date and target visit date unexpectedly small or large? | Some variability in planned and actual visit dates is expected; an excess or lack of such variability may signal fabrication (i.e. a planned ‘4 week follow-up’ occurring a year later, or ‘6-month follow-ups’ consistently occurring at exactly 6 months). |
| 64 | Are there inconsistencies in enrolment dates between IPD, registration record and/or publication? | If enrolment dates are recorded, are they consistent with those reported in associated documents? Fabricated dates may occur before or after the supposed enrolment period. |

**Domain 7 – Inspecting the baseline data (4 checks)**

| **#** | **Check** | **Explanation** |
| --- | --- | --- |
| 65 | Does comparison of baseline data between groups indicate that participants were not randomly allocated? | Alongside more traditional approaches, methods exist for testing for differences in multivariate distributions. These both could be employed to assess whether the distribution of baseline variables systematically differ between study groups, indicating that the data are not consistent with random allocation, or are too similar. |
| 66 | Are only a small number of baseline characteristics collected in IPD? | The proposal is to check that the number of baseline characteristics isn’t under a credible threshold for a genuine trial, which would generally collect/report a large number of variables. |
| 67 | Do statistical tests reveal a difference in variances of baseline variables between groups? | Under true randomization we would expect differences in variances of baseline variables to be compatible with chance 95% of the time. |
| 68 | Is there a difference in kurtosis of baseline variables between groups? | Under true randomization we would expect differences in kurtosis of baseline variables to be compatible with chance. |

Supplementary Figure 1. Outcome of The University of Manchester ethics decision tool

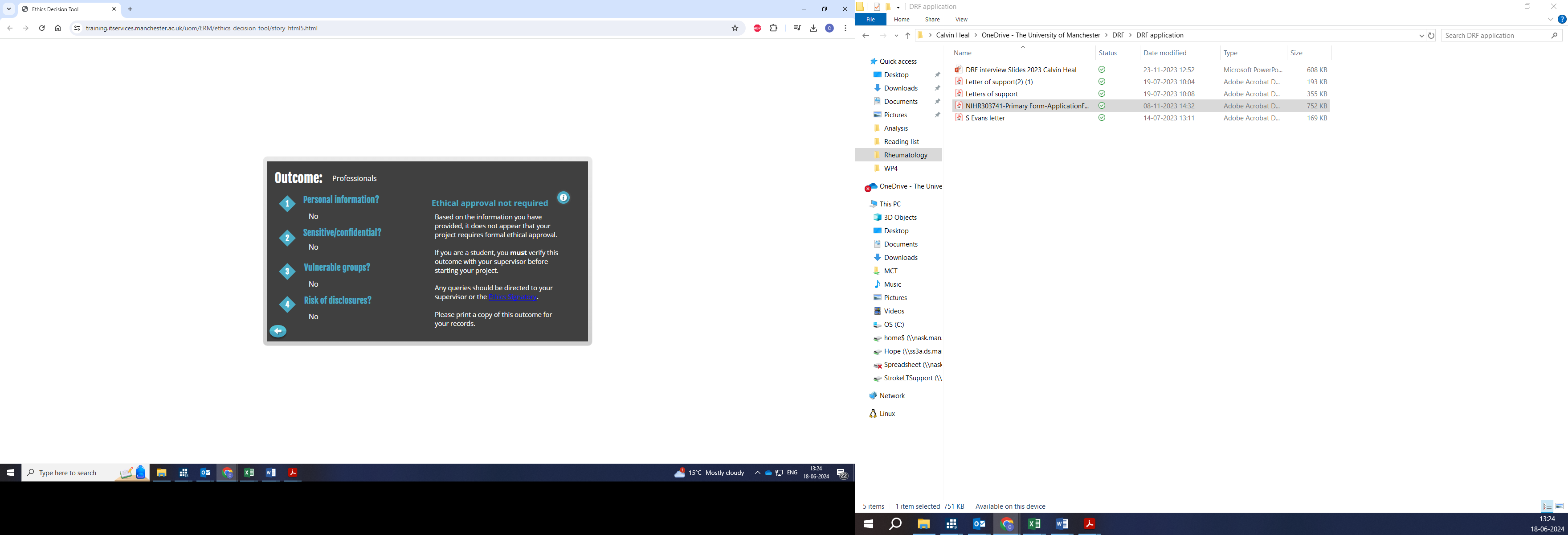

References

1.	Dahlberg JE, Davidian NM. Scientific Forensics: How the Office of Research Integrity can Assist Institutional Investigations of Research Misconduct During Oversight Review. Sci Eng Ethics. 2010 Dec;16(4):713–35.

2.	Lange D, Sahai S, Phillips JM, Lex A. Ferret: Reviewing Tabular Datasets for Manipulation. Computer Graphics Forum. 2023 Jun;42(3):187–98.

3.	Simonsohn U, Nelson L, Simmons J. [109] Data Falsificada (Part 1): ‘Clusterfake’ [Internet]. Data colada. 2023. Available from: https://datacolada.org/109

4.	Carlisle JB. False individual patient data and zombie randomised controlled trials submitted to *Anaesthesia*. Anaesthesia. 2021 Apr;76(4):472–9.

5.	Greenacre M, Öztas Ayhan. Identifying Inliers. 2014 [cited 2025 Feb 6]; Available from: http://rgdoi.net/10.13140/RG.2.1.3651.2406

6.	Wu X, Carlsson M. Detecting data fabrication in clinical trials from cluster analysis perspective. Pharmaceutical Statistics. 2011 May;10(3):257–64.

7.	Venet D, Doffagne E, Burzykowski T, Beckers F, Tellier Y, Genevois-Marlin E, et al. A statistical approach to central monitoring of data quality in clinical trials. Clinical Trials. 2012 Dec;9(6):705–13.

8.	Bordewijk EM, Wang R, Askie LM, Gurrin LC, Thornton JG, Van Wely M, et al. Data integrity of 35 randomised controlled trials in women’ health. European Journal of Obstetrics & Gynecology and Reproductive Biology. 2020 Jun;249:72–83.

9.	Qi S. Use of digital analysis to detect fraud in clinical trials dissertation [Internet]. The John Hopkins University; 2004 [cited 2024 Jul 24]. Available from: https://www.proquest.com/openview/5dcd6d0a68a98bba09994a2bade02c7b/1?pq-origsite=gscholar&cbl=18750&diss=y

10.	Mosimann J, Dahlberg J, Davidian N, Krueger J. Terminal Digits and the Examination of Questioned Data. Accountability in Research. 2002 Apr;9(2):75–92.

11.	Wilkinson J, Heal C, Antoniou GA, Flemyng E, Avenell A, Barbour V, et al. A survey of experts to identify methods to detect problematic studies: stage 1 of the INveStigating ProblEmatic Clinical Trials in Systematic Reviews project. Journal of Clinical Epidemiology. 2024 Nov;175:111512.

12.	Buyse M, George SL, Evans S, Geller NL, Ranstam J, Scherrer B, et al. The role of biostatistics in the prevention, detection and treatment of fraud in clinical trials. Statist Med. 1999 Dec 30;18(24):3435–51.

13.	Brown RJC. The use of Zipf’s law in the screening of analytical data: a step beyond Benford. Analyst. 2007;132(4):344.

14.	Bordewijk EM, Li W, Van Eekelen R, Wang R, Showell M, Mol BW, et al. Methods to assess research misconduct in health-related research: A scoping review. Journal of Clinical Epidemiology. 2021 Aug;136:189–202.

15.	Simonsohn U. [77] Number-Bunching: A New Tool for Forensic Data Analysis [Internet]. Data colada. 2019 [cited 2025 Mar 6]. Available from: https://datacolada.org/77

16.	Grey A, Bolland MJ, Avenell A, Klein AA, Gunsalus CK. Check for publication integrity before misconduct. Nature. 2020 Jan 9;577(7789):167–9.

17.	Burdett S, Stewart LA. A COMPARISON OF THE RESULTS OF CHECKED VERSUS UNCHECKED INDIVIDUAL PATIENT DATA META-ANALYSES. International Journal of Technology Assessment in Health Care. 2002;18(3):619–24.
